# Prior Bariatric Surgery in COVID-19 Positive Patients May Be Protective

**DOI:** 10.1101/2020.12.29.20248991

**Authors:** Megan Jenkins, Gabrielle Maranga, G. Craig Wood, Christopher M. Petrilli, Christine Ren-Fielding

**Affiliations:** NYU Langone Health, 530 1^st^ Avenue, Suite 10S, New York, NY 10016; Geisinger Obesity Institute, 100 North Academy Avenue, Danville, PA 17822; NYU Langone Health, 1 Park Ave, Suite 11-135, New York, NY 10016

## Abstract

**Introduction:** Patients infected with novel COVID-19 virus have a spectrum of illnesses ranging from asymptomatic to death. Data has shown that age, gender and obesity are strongly correlated with poor outcomes in COVID-19 positive patients. Bariatric surgery is the only treatment that provides significant, sustained weight loss in the severely obese. We look at whether prior bariatric surgery correlates with increased risk of hospitalization and outcome severity after COVID-19 infection.

**Methods:** A cross-sectional retrospective analysis of a COVID-19 database from a single, NYC-based, academic institution was conducted. A cohort of COVID-19 positive patients with a history of bariatric surgery (n=124) were matched in a 4:1 ratio to a control cohort of COVID-19 positive patients who were eligible for bariatric surgery (BMI ≥40 kg/m^2^ or BMI ≥35 kg/m^2^ with a comorbidity) (n=496). A comparison of outcomes, including mechanical ventilation requirements and deceased at discharge, was done between cohorts using Chi-square test or Fisher’s exact test. Additionally, overall length of stay and duration of time in ICU were compared using Wilcoxon Rank Sum test. Conditional logistic regression analyses were done to determine both unadjusted (UOR) and adjusted odds ratios (AOR).

**Results:** A total of 620 COVID-19 positive patients were included in this analysis. The categorization of bariatric surgeries included 36% Roux-en-Y Gastric Bypass (RYGB, n=45), 35% laparoscopic adjustable gastric banding (LAGB, n=44), and 28% laparoscopic sleeve gastrectomy (LSG, n=35). The body mass index (BMI) for the bariatric group was 36.1 kg/m^2^ (SD=8.3), which was significantly lower than the control group, 41.4 kg/m^2^ (SD=6.5) (p<0.0001). There was also less burden of diabetes in the bariatric group (32%) compared to the control group (48%) (p=0.0019). Patients with a history of bariatric surgery were less likely to be admitted through the emergency room (UOR=0.39, p=0.0001), less likely to have had a ventilator used during the admission (UOR=0.42, p=0.028), had a shorter length of stay in both the ICU (p=0.033) and overall (UOR=0.44, p=0.0002), and were less likely to be deceased at discharge compared to the control group (OR=0.42, p=0.028).

**Conclusion:** A history of bariatric surgery significantly decreases the risk of emergency room admission, mechanical ventilation, prolonged ICU stay, and death in patients with COVID-19.

## Introduction

The pandemic spread of severe acute respiratory syndrome coronavirus 2 (SARS-CoV-2) and the resulting disease, coronavirus disease-2019 (COVID-19) have been declared a public health emergency of international concern by the World Health Organization (WHO).^1^ As of December 13^th^, 2020, the WHO has reported 70,476,836 cases globally contributing to 1,599,922 deaths.^2^ The United States currently accounts for more than 20% of the global cases and deaths with 15,648,098 cases.^2^

The first documented case in New York City occurred on February 29, 2020, with the city being quickly recognized as the epicenter of the pandemic.^3^ The peak of the first wave occurred on April 6, 2020, with 6,377 confirmed incident cases recorded on one day. As of December 17^th^, 2020, incident daily cases are rising (greater than 700) and cumulatively, 336,567 patients tested positive for COVID-19, with 19,869 confirmed fatalities (5.90%).^4^

Over time, the symptoms of COVID-19 have come into focus, with the most common signs and symptoms being fever, cough, and fatigue, and less common symptoms include headache, hemoptysis, diarrhea, dyspnea, and a decrease of lymphocytes.^5^ More concerning has been the spectrum of outcomes that evolve with COVID-19 infection, ranging from asymptomatic to multi-system organ failure. Several studies have identified risk factors that may lead to poor outcomes with a COVID-19 diagnosis, specifically male gender, age > 60, history of hypertension, type 2 diabetes mellitus, heart disease, kidney damage, and obesity.^4,7-11,25-26^ The association between obesity and mortality is not surprising as there has been a well-documented correlation between obesity and chronic respiratory issues and inflammation.^6^

In a study of New York City patients, Petrilli et al. found that obesity had a high correlation to increased risk of hopsitalization and poor outcomes such as respiratory failure and death.^7^ As obesity is a chronic condition that affects all organ systems, this complex disease results in a multitude of illnesses such as diabetes, hypertension, cardiovascular disease and fatty liver.^8^ Compounding this issue, research has shown that the greater the degree of obesity, the more difficult it is to maintain significant weight loss. Bariatric surgery is the only long-term treatment option for severe obesity that can provide substantial weight loss, which is maintained for years. The use of bariatric surgery as a tool to assist with the reduction or resolution of respiratory symptoms has also been well documented.^9-12^ Since the only long-term treatment option for severe obesity is bariatric surgery, this research aims to further investigate any potential correlation between history of bariatric surgery and improved outcomes in patients diagnosed with COVID-19.

## Methods

### Cohort Selection

A cross-sectional retrospective analysis of a COVID-19 database from a single, NYC-based, academic institution was conducted. This site is also an accredited Center of Excellence through the Metabolic and Bariatric Surgery Accreditation and Quality Improvement Program (MBSAQIP). From the database of tested patients, 8,781 tested positive for COVID-19 and 130 (1.48%) had a history of bariatric surgery. Out of the remaining COVID-19 positive patients without a history of bariatric surgery, a cohort of bariatric surgery eligible patients was created. To be included in the control group, these patients needed a body mass index (BMI) ≥40 kg/m^2^ OR BMI ≥35 kg/m^2^ with a comorbidity (diabetes, hypertension, or hyperlipidemia) present at the time of COVID-19 diagnosis. There were 963 patients that met these inclusion criteria and were eligible to be matched on age (± 3 years) and gender with the bariatric surgery cases. A matching ratio of 4:1 was selected to minimize the loss of cases and to maximize the size of the control group. Of the 130 bariatric cases, there were 4 controls matches found for 124 (95%) of these patients, resulting in a total of 620 subjects used in the analysis (n=124 bariatric cases and n=496 controls). Although the dataset allowed for a 5:1 matching ratio, the number of bariatric cases excluded would increase, resulting in a net loss of statistical power.

### Statistics

The baseline characteristics of the study cohorts were compared using a two-sample t-test for continuous data and Chi-square or Fisher’s exact test for categorical data. Categorical outcomes such as discharge status or deceased at discharge were compared between those with and without a history of bariatric surgery using Chi-square test or Fisher’s exact test. To ensure that the matched case-control design did not have an impact on the results, conditional logistic regression was used to validate the results from the chi-square test results. The overall length of stay and duration of time in ICU were compared between those with and without a history of bariatric surgery using Wilcoxon Rank Sum test. All tests were two-sided and p-values <0.05 were considered significant.

## Results

A total of 620 COVID-19 positive patients were included in this analysis. The categorization of bariatric surgeries included 36% Roux-en-Y Gastric Bypass (RYGB, n=45), 35% laparoscopic adjustable gastric banding (LAGB, n=44), and 28% laparoscopic sleeve gastrectomy (LSG, n=35). The average age was 51.7 years (SD = 12.6) in the bariatric group and 52.1 years (SD = 12.9) in the control group. Both groups were 69% female. There was no statistical difference between these groups as these were the matching criteria. The body mass index (BMI) for the bariatric group was 36.1 kg/m^2^ (SD=8.3), which was significantly lower than the control group, 41.4 kg/m^2^ (SD=6.5) (p<0.0001). There was also less burden of diabetes in the bariatric group (32%) compared to the control group (48%) (p=0.0019). There was no statistical difference in the racial demographics, the week of admission, the distribution of the burden of hypertension, hyperlipidemia, initial inflammatory marker values (CRP and D-dimer), history of myocardial infarction (MI), or history of stroke between the bariatric and control groups (Table 1).

**Table 1.** Bivariate analysis of demographic variables of 620 COVID-19 positive patients with controls age and gender matched to cases with a history of any bariatric surgery

|  | Bariatric Cases<br>n=124 (%) | Bariatric Controls<br>N=496 (%) | p-value |
| --- | --- | --- | --- |
| Variables used in Matching |  |  |  |
| Age, mean ± SD | 51.7±12.6 | 52.1±12.9 | 0.743<sup>1</sup> |
| Female (%) | 69 | 69 | 0.999<sup>2</sup> |
| Variables not used in Matching |  |  |  |
| BMI ± SD | 36.1±8.3 | 41.4±6.5 | <0.0001<sup>1</sup> |
| Race/Ethnicity (%) |  |  |  |
| Spanish/Hispanic | 33 | 34 | 0.799<sup>2</sup> |
| Black | 30 | 29 | 0.930<sup>2</sup> |
| White | 39 | 36 | 0.647<sup>2</sup> |
| Comorbidities (%) |  |  |  |
| Diabetes | 32 | 48 | 0.0019<sup>2</sup> |
| Hypertension | 58 | 66 | 0.085<sup>2</sup> |
| Hyperlipidemia | 44 | 48 | 0.520<sup>2</sup> |
| History of MI | 2 | 3 | 0.778<sup>3</sup> |
| History of stroke | 2 | 6 | 0.121<sup>3</sup> |
| Bariatric Surgery (%) |  |  |  |
| RYGB | 45 (36) | — | — |
| LAGB | 44 (35) | — | — |
| LSG | 35 (28) | — | — |

<sup>1</sup> Two-sample t-test
<sup>2</sup> Chi-square test
<sup>3</sup> Fisher's exact test

Chi-square tests were conducted to determine if there was a comparable difference in the frequency of outcomes between the bariatric and control groups. The following values were confirmed using an unadjusted conditional logistic regression model. When compared to the control group, those with a history of bariatric surgery were less likely to be admitted through the emergency room (UOR=0.39, p=0.0001) and less likely to have had a ventilator used during the admission (UOR=0.42, p=0.028). A Wilcoxon rank sum test showed that the length of stay was longer in the ICU (p=0.033) for those with a history of bariatric surgery. Total length of stay greater than 1 day was less in the bariatric group as well (UOR=0.44, p=0.0002). Overall, ICU admission was lower in the bariatric surgery group, but not significantly so. Finally, those with a history of bariatric surgery were less likely to be deceased at discharge compared to the control group (UOR=0.42, p=0.028). This analysis was conducted again adjusting for BMI, race/ethnicity, diabetes, hypertension, hyperlipidemia, history of MI, and history of stroke. In this model, those with a history of bariatric surgery were still less likely to be admitted from the emergency department (AOR=0.50, p=0.015), however, the remaining outcomes were no longer significant. (Table 2).

**Table 2.** Conditional logistic regression analysis of 620 COVID-19 positive patients with controls age and gender matched to cases with a history of any bariatric surgery

|  | Bariatric Cases<br>n=124 (%) | Bariatric Controls<br>n=496 (%) | Unadjusted Model |  | Adjusted Model* |  |
| --- | --- | --- | --- | --- | --- | --- |
|  |  |  | OR (95% CI) | p-value | OR (95% CI) | p-value |
| Admission Location |  |  |  |  |  |  |
| Outpatient/Other | 61 (49.2) | 54 (31.1) | 1.00 (Ref) | – | 1.00 (Ref) | – |
| Emergency | 63 (50.8) | 342 (69.0) | 0.39 (0.25 – 0.61) | 0.0001<sup>1</sup> | 0.50 (0.29 – 0.88) | 0.015<sup>1</sup> |
| Procedures During Admission |  |  |  |  |  |  |
| Intubation | 2 (1.6) | 18 (3.6) | 0.44 (0.10 – 1.90) | 0.394<sup>2</sup> | – | – |
| Ventilation | 8 (6.5) | 68 (13.7) | 0.42 (0.19 – 0.91) | 0.028<sup>1</sup> | 0.43 (0.18 – 1.04) | 0.060<sup>1</sup> |
| ECMO | 0 (0.0) | 3 (0.6) | – | 0.999<sup>2</sup> | – | – |
| Hemodialysis | 5 (4.0) | 26 (5.2) | 0.76 (0.29 – 2.02) | 0.818<sup>2</sup> | – | – |
| Cardiovascular Events |  |  |  |  |  |  |
| MI | 5 (4.0) | 14 (2.8) | 1.45 (0.51 – 4.10) | 0.558<sup>2</sup> | – | – |
| Stroke | 2 (1.6) | 6 (1.2) | 1.34 (0.27 – 6.72) | 0.664<sup>2</sup> | – | – |
| ICU Admission |  |  |  |  |  |  |
| Any ICU Stay | 10 (8.1) | 73 (14.7) | 0.51 (0.26 – 1.02) | 0.052<sup>1</sup> | 0.61 (0.29 – 1.29) | 0.196<sup>1</sup> |
| Directly to ICU | 6 (4.8) | 44 (8.9) | 0.53 (0.22 – 1.27) | 0.150<sup>1</sup> | 0.73 (0.28-1.89) | 0.516<sup>1</sup> |
| Discharge Disposition |  |  |  |  |  |  |
| Deceased | 8 (6.5) | 68 (13.7) | 0.42 (0.20 – 0.92) | 0.028<sup>1</sup> | 0.51 (0.22 – 1.17) | 0.111<sup>1</sup> |
| ICU Duration |  |  |  |  |  |  |
| 0 Days | 114 (91.9) | 423 (85.3) | – | 0.033<sup>3</sup> |  |  |
| 1-2 Days | 1 (0.8) | 10 (2.0) | – |  |  |  |
| 2-4 Days | 3 (2.4) | 17 (3.4) | – |  |  |  |
| 5-9 Days | 1 (0.8) | 18 (3.6) | – |  |  |  |
| ≥ 10 Days | 5 (4.0) | 28 (5.7) | – |  |  |  |
| Length of Stay |  |  |  |  |  |  |
| ≤ 1 Day | 73 (58.9) | 200 (40.3) | 1.00 (Ref) | – | 1.00 (Ref) | – |
| >1 Day | 51 (41.4) | 296 (59.7) | 0.44 (0.29 – 0.67) | 0.0002<sup>3</sup> | 0.67 (0.41 – 1.09) | 0.105<sup>3</sup> |
| Median (IQR) | 1 (0 – 5) | 3 (0 – 8) | – |  |  |  |

\*Model adjusted for BMI, race/ethnicity, diabetes, hypertension, hyperlipidemia, history of MI, and history of stroke. Results were limited to outcomes that had > 5 events in both groups.

## Discussion

This study demonstrates that bariatric surgery may be protective against severe COVID-19 infection and death for patients with morbid obesity. As mechanical ventilation and length of stay in the ICU have become proxies for severe infection, a history of bariatric surgery improves BMI and is correlated with less severe COVID-19 disease.

To date, the pathophysiology of COVID-19 has not been elucidated; however, it has been shown to cause a wide spectrum of symptoms, from mild upper respiratory tract infection to life-threatening hypoxic respiratory failure. SARS-CoV-2 targets pneumocytes and accentuates the inflammatory response leading to cytokine release and acute respiratory distress syndrome through the viral structural spike protein that binds to the angiotensin-converting enzyme 2 (ACE2) receptor. Severe disease results in hypoxic respiratory failure requiring mechanical ventilation and death.^19^

Many studies have demonstrated that obesity is a risk factor for severe COVID-19.^4,8-11^ The exact mechanism by which obesity results in more severe COVID-19 infection is not completely understood. While several parameters may play a role, the altered respiratory physiology associated with obesity is likely a major contributor. Patients with obesity have decreased functional residual capacity and expiratory reserve volume, as well as ventilation perfusion ratio abnormalities and hypoxemia. These respiratory abnormalities are primarily due to a decrease in chest wall compliance from an accumulation of fat around the ribs, diaphragm, and abdomen.^21^ Nguyen, et al. demonstrated that weight loss after bariatric surgery resulted in a marked improvement in respiratory function. This was demonstrated by extreme improvement in pulmonary function tests as soon as 3 months after bariatric surgery.^17^ This improvement in both restrictive and obstructive respiratory mechanics with weight loss after bariatric surgery may explain the decreased need for mechanical ventilation and severe COVID-19 infection in patients with a history of bariatric surgery.

COVID-19 has a high affinity for ACE2 and has been shown to be the receptor for entry into host cells.^25^ ACE2-expressing tissues are direct targets for SARS-CoV-2, resulting in pathological changes. The expression of ACE2 differs among tissues types. Adipose tissue has been shown to be a one of the human tissues types with the highest expression of ACE2.^26^ While lung tissue has been shown to be a main target tissue affected by SARS-CoV-2, Al-Benna demonstrated that ACE2 expression in adipose tissue is even higher than in lung tissue.^27^ Patients who are obese have more adipose tissue and therefore an increased number of ACE2-expressing cells, possibly leading to an increased susceptibility to COVID-19. As bariatric surgery remains the most effective mechanism of long-term weight loss and therefore overall reduction in adipose tissue, this may be another key mechanism related to the protective effect of bariatric surgery in coronavirus infection.

Furthermore, obesity has been associated with dysregulation of the immune system. Research has demonstrated complex interactions between adipocytes and leukocytes leading to a state of chronic low-grade inflammation with increased levels of inflammatory markers in obesity.^22^ Bariatric surgery results in marked weight loss and improvement in inflammatory markers with reduction in adipose tissue. Many studies have demonstrated COVID-19 as a disease of severe inflammation. D-dimer, a marker for inflammation, is commonly elevated in patients with COVID-19. Yao, et al. showed that D-dimer levels were a reliable marker for in-hospital mortality and they significantly increased with increasing severity of COVID-19.^28^ With weight loss associated with bariatric surgery there is improvement in inflammation, coagulation activation, and fibrinolysis. Ly, et al. showed a significant decrease in D-dimer and CRP activity after bariatric surgery in association with microvesicle-associated tissue factor.^29^ Additionally, significant decreases in pro-inflammatory markers IL-6 and CRP have been shown as early as 12 months after bariatric surgery.^23^ Several studies have demonstrated that lower levels of CRP was associated with less severe COVID-19 disease. Moreover, lower levels of IL-6 was associated with decreased mortality from COVID-19.^24^ Our study did not show a significant decrease in CRP or D-dimer in the post bariatric surgery patients as compared to the control group. However, the substantial weight loss after bariatric surgery may result in overall improvement in immune system function and inflammation and therefore be a contributing factor to the protective effect bariatric surgery has against severe COVID-19 infection.

Our study does have some limitations, such as its retrospective nature. Additionally, it is important to address that our study populations did have a significant difference in BMI. Several studies have shown morbid obesity to be an independent risk factor for critical illness due to COVID-19.^30^ Patients were intentionally not matched based on BMI since the main goal of bariatric surgery is to produce sustained weight loss. Instead, the intention was to evaluate the benefit of bariatric surgery by comparing patients who would qualify for bariatric surgery to their counterparts who had bariatric surgery and the benefit of sustained weight loss. Our data does include an adjusted multivariate analysis including BMI, race/ethnicity, diabetes, hypertension, hyperlipidemia, history of MI, and history of stroke. With this adjustment, a history of bariatric surgery still decreases the need for admission to the hospital independently of BMI. It is important to point out that obesity, as a chronic condition, should be managed with the same algorithm as other chronic conditions. This includes pharmacologic and conservative measures first, followed by procedural interventions if conservative measures are unsuccessful at affecting improvement in the patient. Therefore, in the patients with morbid obesity refractory to conservative measures such as diet, exercise, and pharmacotherapy, bariatric surgery is a logical progression in their care and treatment of their chronic condition.

Furthermore, while our study population is diverse, all of the patients were from a single geographic region. Additionally all patients were treated within a single health system and our outcome assignments might be imperfect as some patients may have been discharged and readmitted elsewhere with critical illness or may have died post-discharge.

Obese patients have been disproportionately impacted by COVID-19 with a higher risk of severe disease and death. Bariatric surgery, which produces a much greater and sustained weight loss when compared to conventional methods, can improve the respiratory and inflammatory factors that likely put this population at increased risk. As the United States has become an epicenter of COVID-19, these findings are even more important, since over 40% of the American population is obese.^20^ In conclusion, our results emphasize the importance of bariatric surgery as a protective factor against severe COVID-19 infection and death in the high-risk obese population and independently decreases the risk of hospitalization.

## Data Availability

Identifiable patient-level data from this project are not available to the public.

